# Comparative analysis of the clinical presentations, cardiovascular and laboratory findings and treatment of heart failure with preserved and reduced ejection fractions in Cameroon: a multicenter cross-sectional study

**DOI:** 10.1101/2020.11.08.20227751

**Authors:** Jérôme Boombhi, Antoin Bele, Mazou N Temgoua, Ba Hamadou, Joel Noutakdie Tochie, Donald Tchapmi, Chris-Nadège Nganou, Liliane Mfeukeu-Kuaté, Alain Menanga, Samuel Kingue

## Abstract

**Background:** Contrarily to past concepts, heart failure with preserved ejection fraction (HFpEF) has become more prevalent than heart failure with reduced ejection fraction (HFrEF). Our objective was to study the clinical, cardiovascular and laboratory findings and therapeutic aspects of HFpEF, compared with those of HFrEF in Yaounde, Cameroon.

**Method:** This was an analytical cross-sectional study carried-out at the Central Hospital, General Hospital and Military Hospital of Yaounde, from January to April 2018 (4 months). 201 patients aged at least 18 years old with an echocardiography confirmed diagnosis of heart failure were been enrolled. We excluded 12 patients because they had a congenital ventricular septal defect (2), chronic cor pulmonale (4), mitral stenosis (5), and pericarditis (1).

**Results:** We found that 45.5% of our patients had HFpEF whereas 37.5% had HFrEF. Patients with HFpEF were older and had a significantly higher incidence of hypertension and obesity. HFrEF was significantly more associated with congestive symptoms than HFpEF. The S3 gallop was significantly more present in patients with HFrEF. Patients with HfpEF had significantly higher rate of atrial fibrillation. Furosemide, spironolactone and digoxin were significantly used more frequently in patients with HFrEF.

**Conclusion:** HFpEF is the most frequent form of heart failure in the hospital setting in Yaoundé, Cameroon. Patients with HFpEF were significantly older and more affected by hypertension and obesity than those with HFrEF. Cardiac ultrasound is indispensable to differentiate between the two entities for better management.

## Introduction

The epidemiological transition of diseases has markedly increased the prevalence of hypertension, diabetes and obesity in low- and middle-income countries located especially in Africa [1,2]. The improvement in the management and surveillance of these cardiovascular risk factors and their complications has led to an increase in life expectancy of patients suffering from these pathologies. During the past two decades, there has been a significant increase in the number of cases of heart failure with preserved ejection fraction (HFpEF) in developed countries, due to a high prevalence of these cardiovascular risk factors and an increment in the ageing population [3-5]. Furthermore, despite several studies done on HFpEF in high-income countries, its diagnosis and management remain an on-going challenges because of the polymorphism of its clinical manifestation [3,4,6] and a myriad of nonconclusive clinical trials respectively [7-12]. There is dearth of data done in Africa on HFpEF. Hence, we aimed to contribute to this knowledge gap by comparing the clinical and cardiovascular and laboratory findings, and the treatment of HFpEF to that of heart Failure with reduced Ejection Fraction (HFrEF).

## Methods

### Study design, settings and participants

This was a cross-sectional analytical study conducted in the cardiology unit of three hospitals, namely: Central, General and Military Hospitals of Yaoundé, Cameroon. It was done over a period of 4 months spanning from January 1, 2018 to April 2018. The study population comprised of all consenting consecutive patients aged 18 years and above with a confirmed diagnosis of heart failure, based on clinical and echocardiographic findings hospitalized or followed-up in one of the three aforementioned hospitals. We excluded all patients with a diagnosis of congenital heart disease, cor pulmonale, mitral stenosis or pericarditis.

#### Operational terms

Heart Failure with reduced Ejection Fraction (HFrEF) was defined as the presence of signs and/or symptoms of heart failure with a left ventricular ejection fraction (LVEF) <40%. Heart Failure with preserved Ejection Fraction (HFpEF) was defined as the presence of signs and/or symptoms of heart failure, a LVEF > 50%, a structural heart anomaly (left ventricular hypertrophy and/or left atrial dilation) and/or diastolic dysfunction. A diastolic dysfunction was considered present if: the ratio *E/e′* ≥ 13 (ratio between the peak velocity flow E through the mitral valve on pulse wave Doppler heart ultrasound and the velocity *e*′ at early diastole of the flow at the mitral valve on Doppler heart ultrasound) and/or *e*′ < 9cm/s. Left ventricular hypertrophy was defined as a left ventricular mass ≥115g/m^2^ in men and ≥95g/m^2^ in women. Left atrial dilation was defined as a left atrial volume >34 ml/m^2^ or a surface area >20cm^2^. The signs and symptoms of heart failure considered were those given by the European Society of Cardiology 2016 (ESC 2016) [13].

#### Data collection

Data were collected from registries of hospitalization and consultation, from patients’ medical records, and directly from patients or their families through by interview-administration of pretested structured questionnaire. Data were recorded using data entry sheets. Variables studied were: sociodemographic (age and gander), past history (hypertension, diabetes mellitus, tobacco abuse, obesity and/or overweight, dyslipidemia, chronic alcoholism, sedentary lifestyle, chronic kidney disease, cerebrovascular diseases), physical findings (blood pressure, pulse, respiratory rate, body mass index, signs and symptoms of heart failure) and treatment modalities.

#### Statistical analysis

All data collected were entered using CSPro 7.0. Analysis were done using SPSS version 20.0. Qualitative variables were expressed as numbers and percentages, while quantitative variables were expressed as means and standard deviation. The chi-square test was used to find associations between dependent and categorical variables while the student t-test was used to find associations between continuous variables.

## Results

A total of 201 patients with heart failure were enrolled in the study. We excluded 12 (5.97%) patients for the following reasons: two had a congenital ventricular septal defect, four had chronic cor pulmonale, five had mitral stenosis and one had pericarditis. Eighty-six (45.50%) patients had HFpEF while 71 (37.57%) had HFrER. The remaining 32 (16.93%) patients had an intermediate ejection fraction.

### Comparison of the clinical findings in HFpEF and HFrEF

The mean age of our study population was 65.69±13.58 years. Patients with HFpEF were significantly older (p = 0.003), had hypertension (p= 0.034) and obesity (p = 0.013). There were also more patients of the female sex, with diabetes, tobacco abuse and dyslipidemia in the group with HFpEF, but these differences were not significant **(Table 1)**. HFpEF was more of sudden onset while HFrEF had a more insidious onset and more congestive signs/symptoms **(Table 2)**. Mean systolic blood pressure was significantly higher in patients with HFpEF (p = 0.020) while the heart rate was significantly higher in patients with HFrEF (p < 0.001). The S3 galop sound was significantly more present in patients with HFrEF while there was no significant difference in the occurrence of the S4 galop sound in the two groups (p=0.376) **(Table 2)**. Hypertension was identified as the cause of heart failure mainly in patients with HFpEF (p<0.001) **(Table 3)**.

**Table 1:** Cardiovascular risk factors, past history.

| Parameters | HFpEF n(%) | HFrEF n(%) | p value |
| --- | --- | --- | --- |
| Age (mean ± SD) | 70,90±11,88 | 59,38±12,87 | **0,003** |
| Female sex | 49(56,98) | 35(49,30) | 0,081 |
| Hypertension | 67(77,91) | 45(63,38) | **0,034** |
| Type 2 diabetes | 31(36,05) | 22(30,99) | 0,310 |
| Tobacco smoking | 13(15,12) | 9(12,68) | 0,409 |
| Dyslipidemia | 43(50,00) | 25(35,29) | 0,187 |
| BMI ≥25 | 64(74,42) | 40(56,34) | **0,013** |
| sedentarity | 58(67,44) | 40(56,34) | 0,103 |
| Alcohol intake | 26(30,23) | 28(39,44) | 0,149 |
| Chronic kidney disease | 5(5,81) | 2(2,82) | 0,308 |
| Cerebrovascular disease | 5(5,81) | 3(4,23) | 0,470 |
| hyperthyroidism | 2(2,32) | 2(2,82) | 0,615 |

**Table 2:**
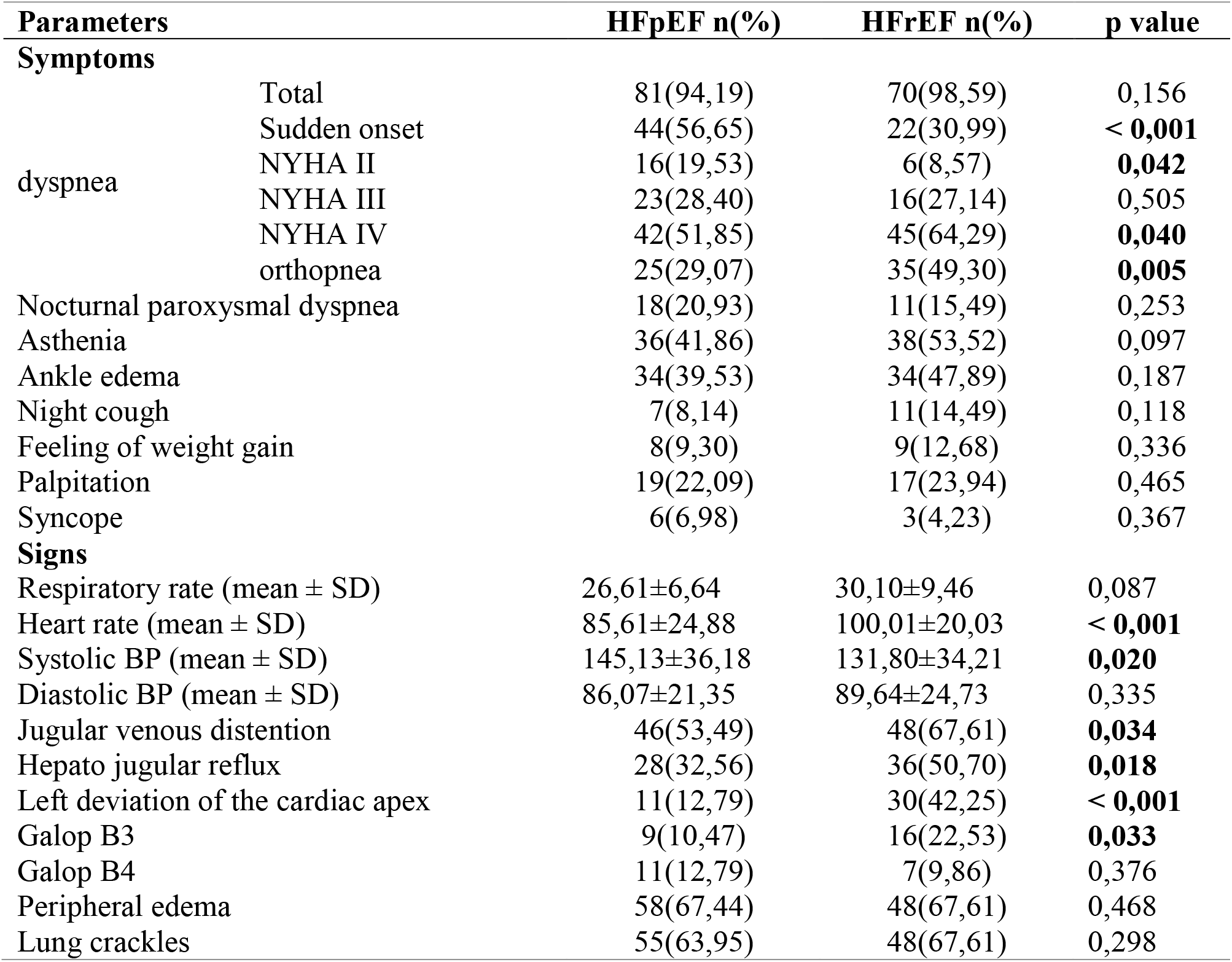
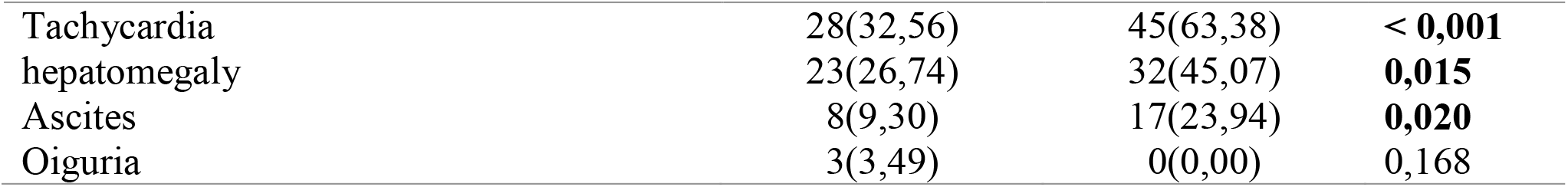
signs and symptoms of heart failure.

**Table 3:**
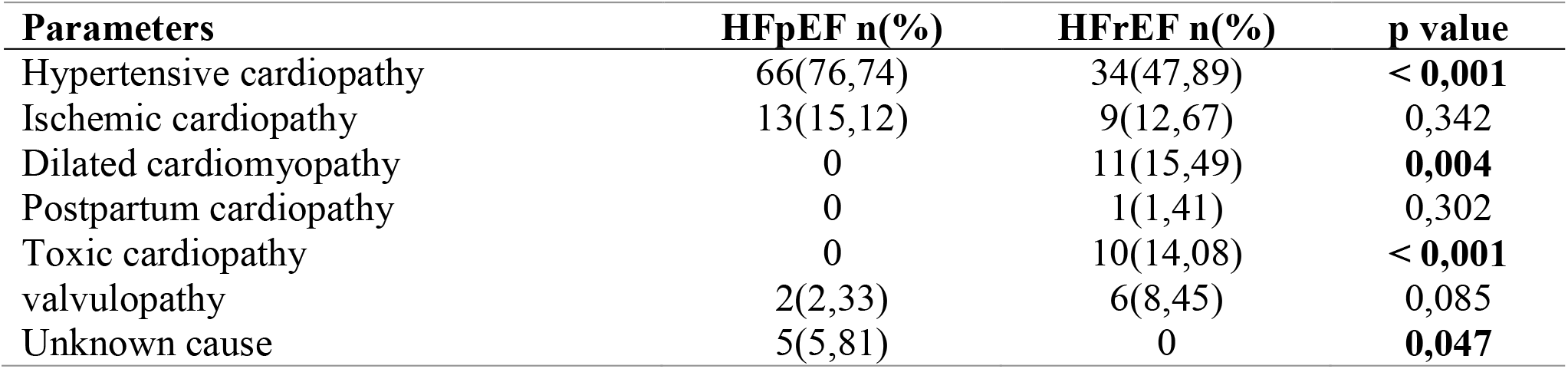
causes of heart failure.

### Comparison of paraclinical findings

#### Electrocardiographic findings

Atrial fibrillation was more frequent in patients with HFpEF (p = 0.013) and more ventricular extra systoles were occurred in the patients with HFrEF (p = 0.036) **(Figure 1)**.

**Figure 1:**
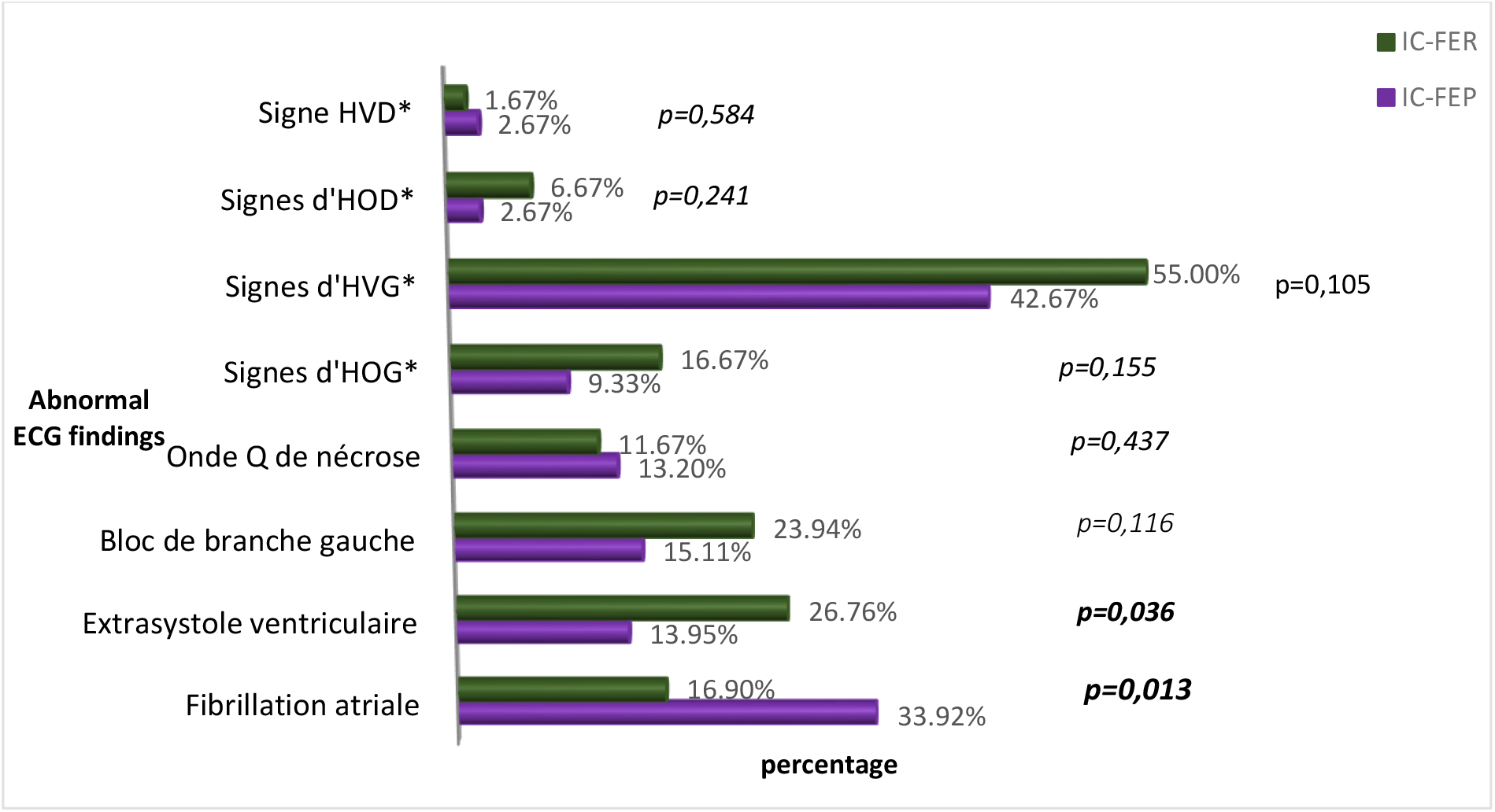
electrocardiographic findings in the two groups. *HOG : Left atrial enlargement, HVG : left ventricular hypertrophy, HOD : right atrial enlargement, HVD : right ventricular hypertrophy.

### Cardiac transthoracic doppler ultrasound findings

The mean mass of the left ventricle was more important in patients with HFpEF (p = 0.004), while the mean area of the left atrium was higher in patients with HFrEF (p = 0.010) **(Table 4)**. Left ventricular hypertrophy (LVH) was more frequent in patients with HFpEF (p = 0.001) **(Figure 2); i**t was more concentric in patients with HFpEF (p < 0.001) and more eccentric in those with HFrEF (p = 0.006) **(Figure 3)**. Abnormal relaxation of the right ventricle was more frequent in patients with HFpEF while patients with HFrEF had a more restrictive profile of the mitral valve **(Figure 4)**.

**Table 4:** Distribution of echocardiographic findings in the two groups.

| **Parameters** | **HFpEF (%)** |  | **HFrEF (%)** |  | **p** |
| --- | --- | --- | --- | --- | --- |
|  | Mean | SD | Mean | SD |  |
| Surface of the RA(cm<sup>2</sup>) | 21,48 | 14,88 | 21,73 | 5,38 | 0,074 |
| Surface of the LA (cm<sup>2</sup>) | 24,41 | 9,91 | 28,38 | 6,38 | **0,010** |
| LV mass index (g/m<sup>2</sup>) | 120,20 | 29,46 | 107,10 | 25,56 | **0,004** |
| TDDL V (mm) | 48,50 | 7,83 | 63,47 | 8,12 | **<0,001** |
| TSDL V (mm) | 31,89 | 8,09 | 54,36 | 7,85 | **<0,001** |
| Valve leaflet retraction (%) | 35,30 | 8,37 | 14,89 | 4,93 | **<0,001** |
| Ejection fraction (Teicholz) (%) | 63,93 | 9,33 | 27,65 | 7,19 | **<0,001** |
| SPAP (mmHg) | 44,13 | 19,01 | 53,69 | 17,89 | **0,010** |

*RA : right atrium, LA : left atrium, LV : left ventricle, TDDL V : telediastolic diameter of the left ventricle, TSDL V : telesystolic diameter of the left ventricle, SPAP : systolic pulmonary arterial pressure.*

**Figure 2:**
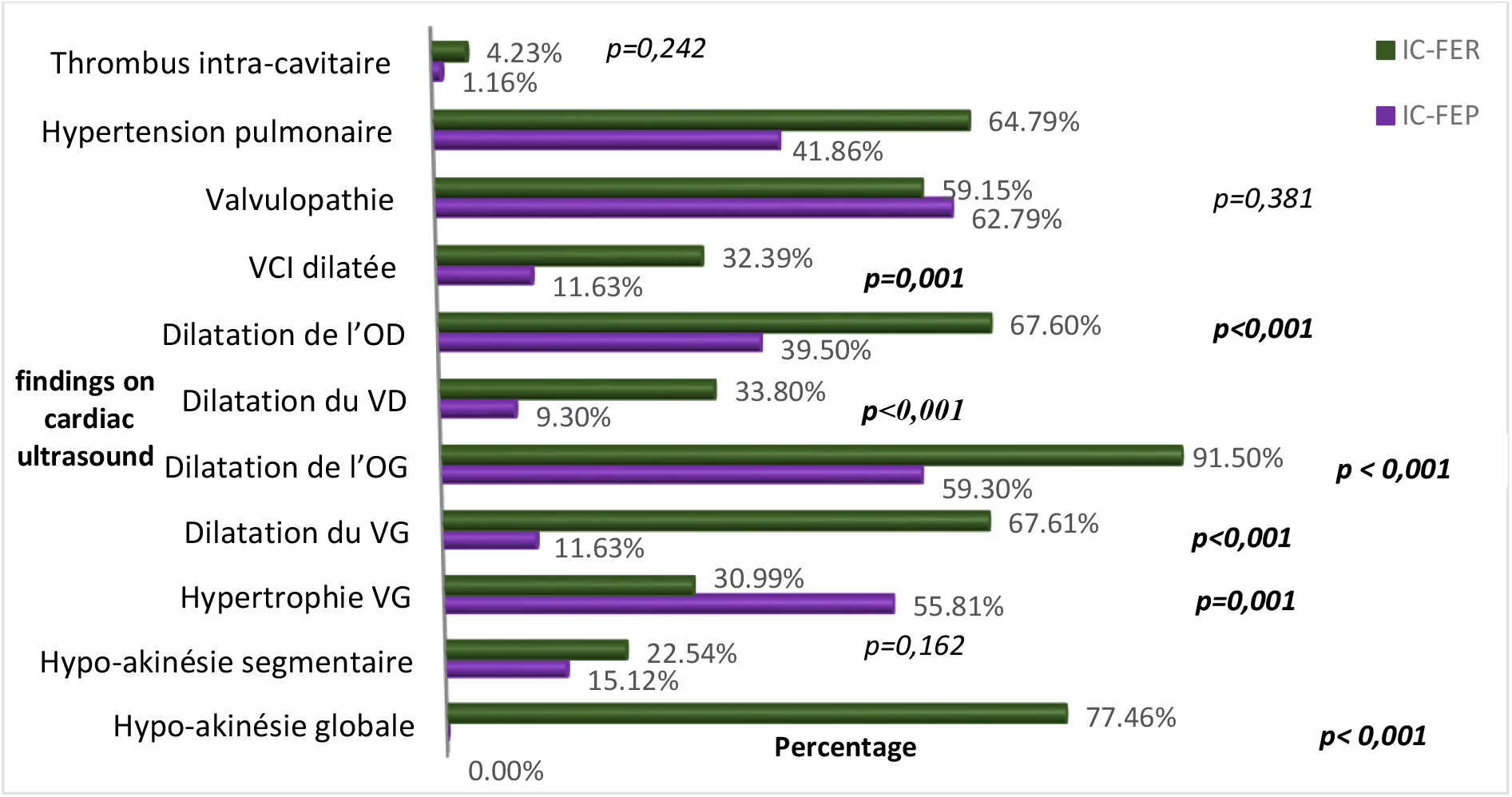
echocardiographic findings in the two groups. *\*VG : left ventricle, OG : left atrium, VD : right ventricle, OD : right atrium, VCI : inferior vena cava*

**Figure 3:**
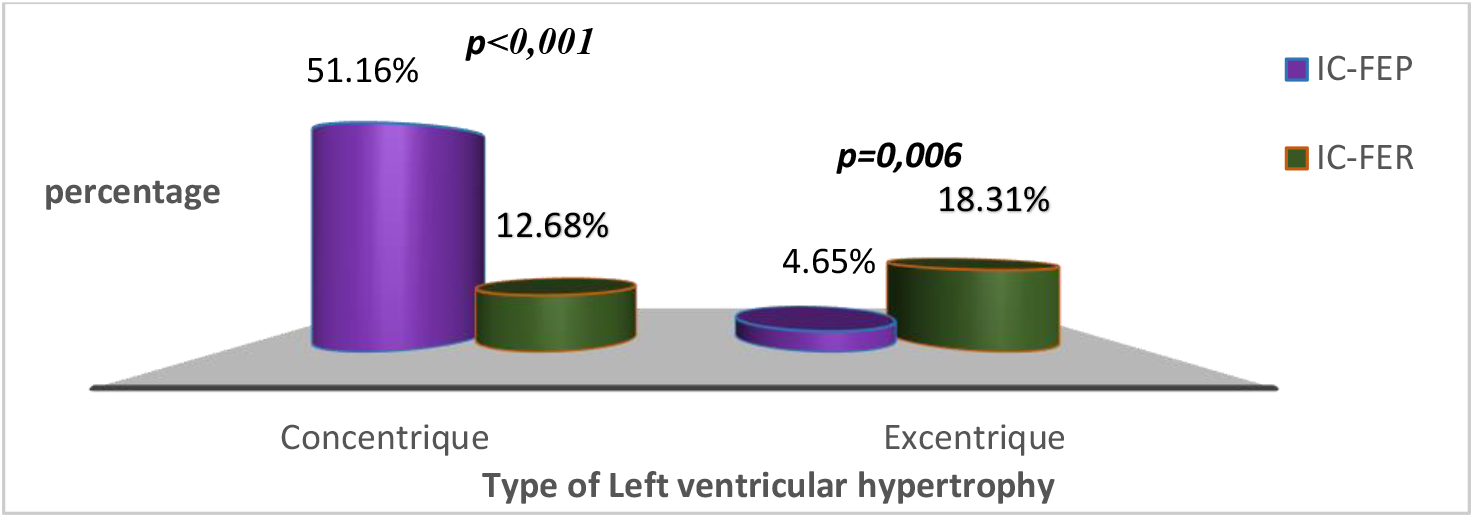
Distribution of patients according to the type of left ventricular hypertrophy.

**Figure 4:**
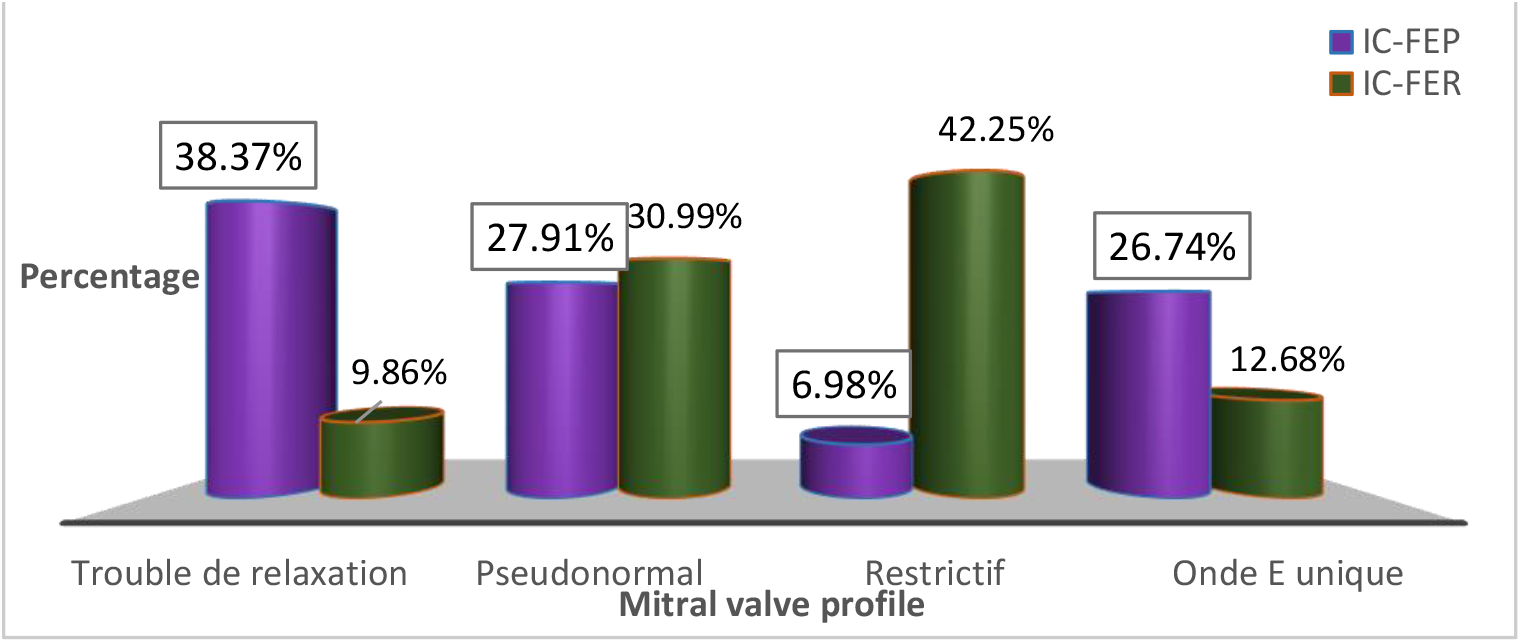
Distribution of patients according to the mitral valve profile.

### Radiologic findings

Cardiomegaly was significantly more prevalent in patients with HFrEF (p<0.001) **(Table 5)**.

**Table 5:** radiologic findings.

| **Parameters** | **HFpEF(N\*=40)** | **HFrEF(N\*=39)** | **p value** |
| --- | --- | --- | --- |
| Cardiac Index (mean ± SD) | 0,53±0,05 | 0,61±0,06 | **<0,001** |
| Cardiomegaly (%) | 31(77,50) | 37(94,87) | **0,026** |
| Pulmonary congestion (%) | 35(87,50) | 34(87,18) | 0,615 |
| Pleural effusion (%) | 7(17,50) | 13(33,33) | 0,118 |

N\*= number of patients with a chest X ray

### Laboratory findings

There was no significant difference in hemoglobin level, estimated glomerular filtration rate (eGFR) and low-density lipoproteins (LDL) cholesterol between the two groups **(Table 6)**. Anemia was more prevalent in patients with HFpEF and altered kidney function was found more in patients with HFrEF, but these differences were not significant **(Figure 5)**.

**Table 6:** laboratory findings.

| **Parameters** | **HFpEF** |  | **HFrEF** |  | **p value** |
| --- | --- | --- | --- | --- | --- |
|  | **Mean** | **SD** | **Mean** | **SD** |  |
| Hb (g/l) | 12,07 | 3,18 | 12,16 | 2,08 | 0,848 |
| Blood urea nitrogen (g/l) | 0,49 | 0,34 | 0,57 | 0,42 | 0,519 |
| Serum creatinine (mg/l) | 17,95 | 20,65 | 16,18 | 9,31 | 0,075 |
| GFR (ml/min/1,73 m<sup>2</sup>) | 67,08 | 33,92 | 62,00 | 25,35 | 0,321 |
| Total cholesterol ( g/l) | 1,77 | 0,50 | 1,67 | 0,54 | 0,495 |
| triglycerides (g/l) | 1,00 | 0,42 | 0,88 | 0,40 | 0,333 |
| LDL cholesterol (g/l) | 1,02 | 0,42 | 0,90 | 0,40 | 0,308 |
| HDL cholesterol (g/l) | 0,62 | 0,63 | 0,60 | 0,40 | 0,589 |

\*HOG : Left atrial enlargement, HVG : left ventricular hypertrophy, HOD : right atrial enlargement, HVD : right ventricular hypertrophy.

**Figure 5:**
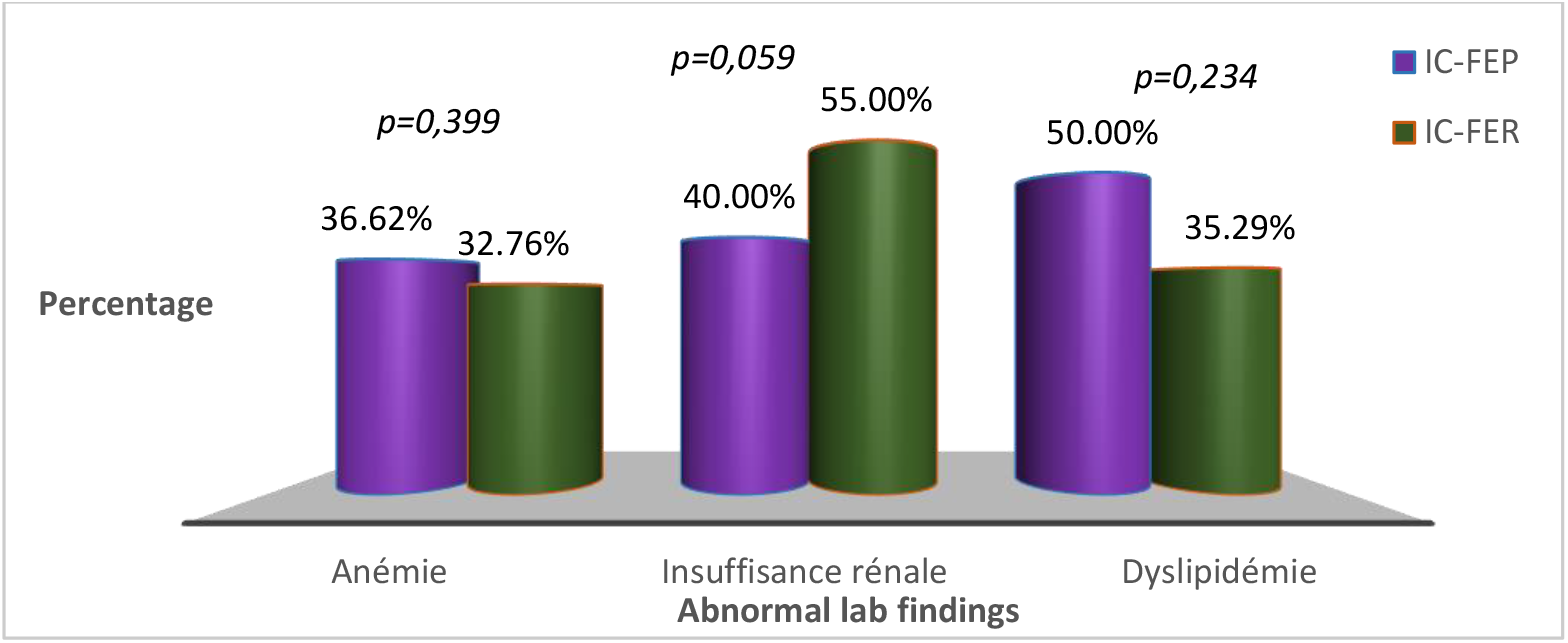
Prevalence of anemia, kidney injury and dyslipidemia in the two group of patients.

### Treatment

Furosemide, spironolactone and digoxin were used more frequently in patients with HFrEF (p<0.05). Angiotensin Converting Enzyme Inhibitors (ACEI), Angiotensin Receptor Blocker (ARB) and beta blockers were also used more frequently in patients with HFrEF but the difference was not statistically significant **(Figure 6)**.

**Figure 6:**
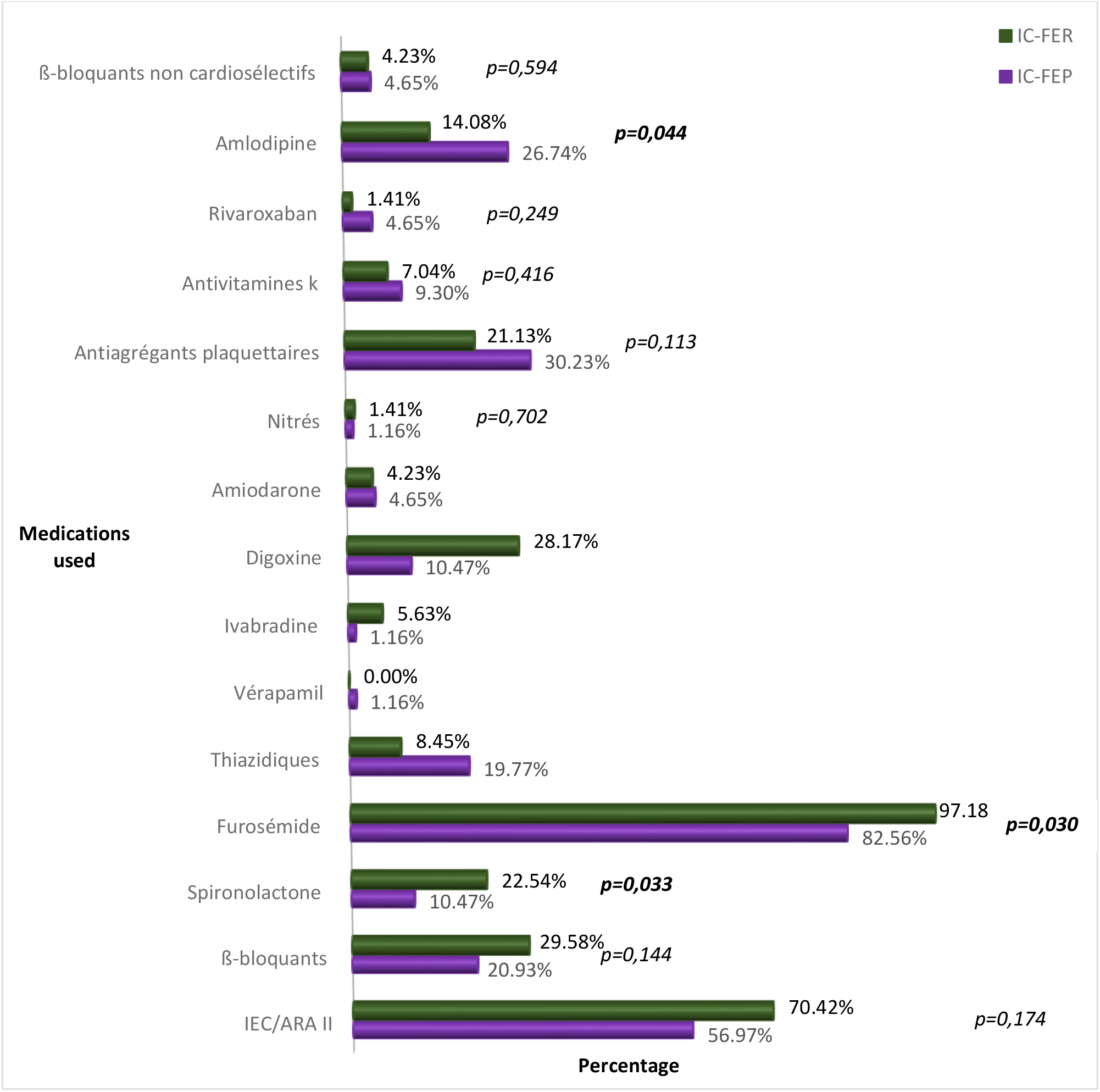
Medications used for treatment of heart failure in the two groups.

## Discussion

The current multicenter study conducted in three major referral cardiology units of Cameroon aimed at comparing the clinical presentations, cardiovascular investigations/laboratory findings and treatment of HFpEF and HFrEF. We enrolled 201 patients with heart failure among which 45.5% had HFpEF and 37.5% HFrEF. Patients with HFpEF were older and had a significantly higher incidence of hypertension and obesity. Patients with HFrEF had more signs of fluid overload than those with HFpEF. The S3 gallop was significantly more present in HF-rEF. Patients with HFpEF had significantly higher rate of atrial fibrillation. Furosemide, spironolactone and digoxin were significantly more frequent in patients with HFrEF.

The prevalence of HFpEF (45.50%) in our study is comparable to those reported in Western countries which varies between 24 to 73% depending on the diagnostic criteria used for HFpEF [6,14-16]. This prevalence is largely above 10% found by Kingue *et al*. in 2005 in a hospital based study on 167 patients with heart failure in Yaounde, Cameroon [17] and that of Mboup *et al*.[18] in a hospital based study on 32 patients in Dakar, Senegal in 2013. The differences observed are linked to the methodology used. We did a cross-sectional analytical study, including patients who were either in acute phase or those with chronic heart failure already on treatment; meanwhile, according to reports from France [19], the ejection fraction tends to increase after the patient is discharged from the hospital and/or placed on treatment, hence, there is a possibility of overestimation of the prevalence of HFpEF in the present study. Also, approximately 80 million people had hypertension in sub-Saharan Africa in the year 2000, and this figure is expected to double by the year 2025 [20]. This implies that the prevalence of HFpEF will also be on a rise since hypertension is one of the main determinants of this pathology [14,21,22].

Patients with HFpEF were significantly older than patients with HFrEF (p=0.003). This finding is similar to what is described in developed countries were patients with HFpEF tend to be older (69 to 78 years) compared to patients with HFrEF (56 to 74 years) [6,23,24]. The patients in our study were older than patients in the study done by Mboup *et al*. in Dakar [18] where the mean age was 65.7 years. The number of female patients were higher in the group of HFpEF, and our results confirmed the fact that more women have HFpEF (42-85%) and more men have HFrEF (20-65%) [3,4,16,25,26]. The fact that this difference was not statistically significant is probably due to a small sample size. Hypertension and obesity were significantly higher in patients with HFpEF. Our results are similar to studies done in developing countries where hypertension and obesity were significantly more present in patients with HFpEF [3,4,24,25,27,28]. Type 2 diabetes mellitus, tobacco abuse and dyslipidemia were also more frequent in patients with HFpEF but the difference was not statistically significant. These results are different from studies done in Western countries in which these factors were found to be more associated with HFrEF [4,16,24,25,28,29].

Dyspnea was the major presenting complain in both group of patients but was more frequent in patients with HFrEF. Bhatia *et al*. [4], Maestre *et al*. [30], and Chinali *et al*. [28] had similar results. The dyspnea was of sudden onset mostly in patients with HFpEF, this can be explained by the mode of evolution of the disease which usually presents with “flash” pulmonary edema, in pauci or asymptomatic patients, due to a precipitating factor [31]. More than half of the patients in both groups were at stage IV of NYHA, but it is more frequent in patients with HFrEF. The fact that most patients were at stage IV illustrates the severity of heart failure in Africa as shown by studies done by Kingue *et al*. in 2005, where 53% of the patients were at stage III or IV [17]. Orthopnea was higher in patients with HFrEF (*p=0,005*). Likewise, Chinali *et al*. [28], Bhatia *et al*. [4] and Gupta *et al*. [16] found a higher prevalence of orthopnea in patients with HFrEF but it was not statistically significant. Mean systolic blood pressure was higher in HFpEF while heart rate was higher in patients with HFrEF. Similar findings were reported in studies from high-income countries [24–27,29,32]. Jugular venous distention, hepato-jugular reflux, hepatomegaly, and ascites were more frequent in patients with HFrEF. These results are similar to those found in Europe and the US [4,25,27,30]. In several studies notably those of Bhatia *et al*. [4], Yancy *et al*. [25], Fanarow *et al*. [27], and Bishu *et al*. [32], the occurrence of ankle edema was significantly higher in patients with HFpEF ; but in our study, it was not significantly higher in patients with HFrEF (p=0.468). The S3 galop was predominant in patients with HFrEF (p=0.033) while the S4 galop was predominant in patients with HFpEF (p=0.376). Bhatia *et al*. also had more S3 galop in patients with HFrEF but less S4 galop in patients with HFpEF [4]. Also, in studies done by Yancy *et al*. [25], Fanarow *et al*. [27], and Bishu *et al*. [32], pulmonary crackles were more frequent in patients with HFpEF; but in our study, it was more frequent in patients with HFrEF though not significant (p=0,298).

Arterial hypertension was the major cause of heart failure in both groups but it was more implicated in the development of HFpEF [33]. Our study shows that hypertension remains the leading cause of heart failure in our milieu and confirms the major role it plays in the pathogenesis of HFpEF as described in the literature [4,6,31].

On ECG, atrial fibrillation was more prevalent in patients with HFpEF, while ventricular ectopic beats were more frequents in patients with HFrEF. Left bundle branch block was more frequent in patients with HFrEF and pathologic Q waves in patients in HFpEF, but these differences were not significant. Our results with the exception of the distribution of pathologic Q waves in the two groups are similar to those described in the literature; a study done by Maestre *et al*. found a higher prevalence of pathologic Q waves in patients with HFrEF. Indeed, Maestre *et al*. found in their studies that a left bundle branch bloc and a pathologic Q wave occurred respectively in 27.9% and 18.6% of patients with HFrEF against 13.7% and 6.3% HFpEF (p=0.037 and 0.032 respectively) [30]. On cardiac ultrasound, concentric left ventricular hypertrophy was more frequent in patients with HFpEF, while eccentric left ventricular hypertrophy was significantly higher in patients with HFrEF. This pattern is similar to what is described in the pathophysiology of HFpEF and HFrEF, whose causes are hypertension [21,23] and myocardial dysfunction [34] respectively. Cardiomegaly was found to be more prevalent in patients with HFrEF. The proportion of pleural effusion was similar in both groups. Our data were similar to the classical findings described in the literature [31,35].

The group of patients with HFpEF had a higher proportion of anaemia (36.62% *vs*. 32.72%, *p=0,399*) while decreased kidney function was found in higher proportion in patients with HFrEF (40% *vs*. 55%, *p=0,059*). Distribution of anemia in our study was similar to that observed by Maestre *et al*. [30] and by Steinberg *et al*. [26]. Ather *et al*., in a cohort evaluating the effect of non-cardiovascular comorbidities on HFpEF and HFrEF, found a significantly higher proportion of anemia in patients with HFpEF [36]. The distribution of altered kidney function in our study was also similar to that found in other studies. Yancy *et al*. [25] found a similar proportion of kidney injury in both groups (26% against 26%, *p=0,98*), and Chinali *et al*. [28] found a non-significant higher proportion in patients with HFrEF (60% *vs*. 64%, *p=0,11*).

Furosemide, spironolactone and digoxin were mostly used in patients with HFrEF (p<0,05); ACEI/ARB and beta blockers were also used more frequently in this group but the difference was not statistically significant. Similar findings are reported in studies from developed countries [24-27,29,32]. Our small sample size might be the reason why there was no significant difference in the usage of ACEI/ARB and beta blockers between the two groups.

## Conclusion

HFpEF is the most frequent form of heart failure in the hospital setting in Yaoundé. Patients with HFpEF were significantly older and more affected by hypertension and obesity than those with HFrEF. Cardiac ultrasound is indispensable to differentiate between the two entities for better management.

### What is already known on this topic

Heart failure with preserved ejection fraction (HFpEF) is the most frequent form of heart failure. A clear clinical picture has been described in developed countries. This entity concerned patients with several comorbidities like: diabetes, obesity and hypertension. There is a challenge in management of these patients with not well defined therapeutic measures apart symptomatic treatment and management of comorbities

### What this study adds

In Sub Saharan Africa, clinical characteristics of the patients could be the same like abroad with a significantly predominance in older patient, hypertensive and obese. Treatment of these conditions is slightly different from those with HFrEF because in this group patients presented more with congestive heart failure and atrial fibrillation. Most studies are needed to compare outcomes of both groups in our context.

## Data Availability

Data could be shared after reasonable request to the corresponding author

## Author’s contribution

Study conception, data collection, analysis and interpretation of the results: JB, AB, BH, CNN, LMK

Critical revision: MNT, DT

Supervision of all steps: AM, SK

All authors approved the final version of the paper

## Acknowledgements

We acknowledge all participants who accepted to contribute in this study; also the administrative authorities of all setting where we have recruited patients

